# What is the impact of high-profile end-of-life disputes on Paediatric Intensive Care (PIC) trainees? Original research

**DOI:** 10.1101/2022.10.23.22281413

**Authors:** Clare Bell, Mariana Dittborn, Joe Brierley

## Abstract

**Introduction:** This study explores UK Paediatric Intensive Care (PIC) trainees’ thoughts and feelings about high-profile end-of-life cases recently featured in the press and social media and the impact on their career intentions.

**Methods:** Semi-structured interviews were conducted with nine PIC-GRID trainees (Apr-Aug 2021). Interview transcripts were analysed using thematic analysis.

**Results:** Six main themes were identified: (i) All participants wished to do what was best for the child, feeling conflicted if this meant disagreeing with parents. (ii) Interviewees felt unprepared and expressed deep concern about the effect of high-profile cases on their future career. (iii) They highlighted too often being shielded from involvement in challenging discussions. (iv) Working in a supportive environment is crucial, underscoring the importance of clear and unified team communication, but specific training on the ethical and legal nuances of such cases is required. (vi) All had purposefully minimised their social media presence. (vi) All had reconsidered training in PIC due to concerns about future high-profile end-of-life disputes; despite describing this as a cause of concern and anxiety, all were still in training.

**Conclusion:** UK PIC trainees feel unprepared and anxious about future high-profile cases. A parallel can be drawn to child protection improvements following significant educational investment after Government reports into preventable child abuse deaths. Models for supporting trainees and establishing formal PIC training are required to improve trainees’ confidence and skills in managing high-profile cases. Further research with other professional groups, the families involved, and other stakeholders would provide a more rounded picture.

- **What is already known on this topic** – PIC trainees report high levels of distress associated to children’s death and disagreements with the families.
- **What this study adds** – PIC doctors are extremely concerned about future high-profile end-of-life disputes, the effect on themselves and their career. They feel unprepared to manage them and require more training, experience and recognition of this.
- **How this study might affect research, practice or policy** – PIC trainee curriculum and training scheme should prepare doctors to deal with high-profile end-of-life cases to ensure children and families receive appropriate care and avoid PIC-induced moral injury and workforce burdens.

## Introduction

End-of-life decisions in children are intensely emotive, especially for the child and family. In Paediatric Intensive Care (PIC), such choices include decisions to limit life-sustaining therapies (LST), making PIC a stressful and emotionally draining speciality. Given the increasing number of children living with complexity or life-limiting conditions, it is unsurprising that the small number of PIC cases in which courts are approached to determine children’s best interests has also increased. The UK Paediatric Critical Care Society identified staff mental health and stress as the most crucial topic requiring further research.(1) PIC-moral distress arises from continuing invasive treatment perceived not to be in the children’s best interests or harmful.(2) By the time disagreements are resolved, including via the courts, PIC teams have provided non-medically indicated LST long past a time they thought it helpful. Consequently, high levels of emotional distress and burnout are seen in PIC staff after a child’s death or following conflict with parents.(3)

Recently several high-profile cases involving the High Court have featured in the national and international media and caused social media storms.(4–6) Resentment and negativity quickly focused on hospitals, often directly targeting individual PIC doctors. Sometimes this led to protests outside hospitals and death threats to staff. PIC trainees anecdotally described these cases as a genuine and growing concern, particularly the press and social media involvement. Some suggest such cases led them to reconsider a PIC career.

Given current workforce pressures, such views threaten to have a long-term negative impact on recruitment and retention,(7) worsening the effects of PIC-induced moral injury and workforce burdens.(8) However, a gap exists in our understanding of the impact of this on PIC trainees, tomorrow’s PIC consultants. We elicited PIC trainees’ thoughts and feelings about high-profile end-of-life cases, how their training might usefully be adapted (if necessary) and the potential impact on career intentions.

## Methods

### Study design

A qualitative study of PIC trainees based on semi-structured interviews in which they express their thoughts and experiences around the impact of high-profile PIC-end-of-life disputes.(9)

### Setting, participants and sample size

Purposeful sampling targeted PIC-GRID trainees, who provide most future PIC consultants. Eligibility criteria: UK PIC-GRID Trainee. Exclusions: non-PIC GRID trainees, e.g., general paediatric trainees, anaesthetists, junior doctors not training, adult ICU trainees and overseas doctors as fewer become PIC consultants.

PIC GRID trainees attending a national trainee day received an email containing the participant information leaflet and opt-in reply. A sample size of 6-10 participants was anticipated likely for data saturation, generating a rich and manageable data set.

### Data collection methods

The authors developed an interview topic guide, ensuring the interviewer could adapt to explore any deeper feelings arising during the interview. They trialled it before the study and made it available to participants before the interview. CB conducted the interviews alone at her workplace in a quiet room, following written consent at a mutually convenient time over MS Teams© (April-August 2021).

Interviews lasted 30-90 minutes and were audio recorded. Afterwards, participants were offered information about emotional support and contacted the next day to enquire how they were coping post-interview.

### Data analysis

Interviews were transcribed verbatim and anonymised using alphanumeric codes (I1, I2…). Transcripts were analysed using NVivo® following Braun and Clarke’s six-step method for thematic analysis.(10) Inductive coding of transcriptions was performed by CB and checked by JB and HB. Themes identified were discussed within the research team until agreement on the codes and themes most accurately representing the original data was reached. No themes were decided in advance, all arose from the data. Representative quotes were selected for each theme to illustrate the essential points of interpretations. CB kept a research journal, allowing the research team to look back to check whether initial feelings had impacted the thematic analysis and to question any evident bias in the initial feelings post each interview.

### Reflexivity

CB (MBBS, BSc, MSc) being a female final year UK General Paediatric Trainee, ensured commonality between paediatric doctors allowing trainees to feel at ease and has elicited a rich data set (31), which may not have been yielded if trainees had to explain the nuances of medicine and PIC to the researcher. (30)

### Ethical considerations

Study approval by GOSH-Clinical Research Adoptions Committee (ID 20HL11), UCL-ethics committee (ID 18271/001), and the Health Research Authority Research Ethics Committee (ID 278437).

### Funding

No funding.

## Results

### Participants’ characteristics

Ten trainees opted in, 1 did not fulfil the eligibility criteria as was an Adult Anaesthetic Trainee, nine trainees were interviewed, aged 30-50 years, with interview data organised into six major themes. Representative quotes for each theme are provided in Tables 3, 4 and 5.

**Table 1.**
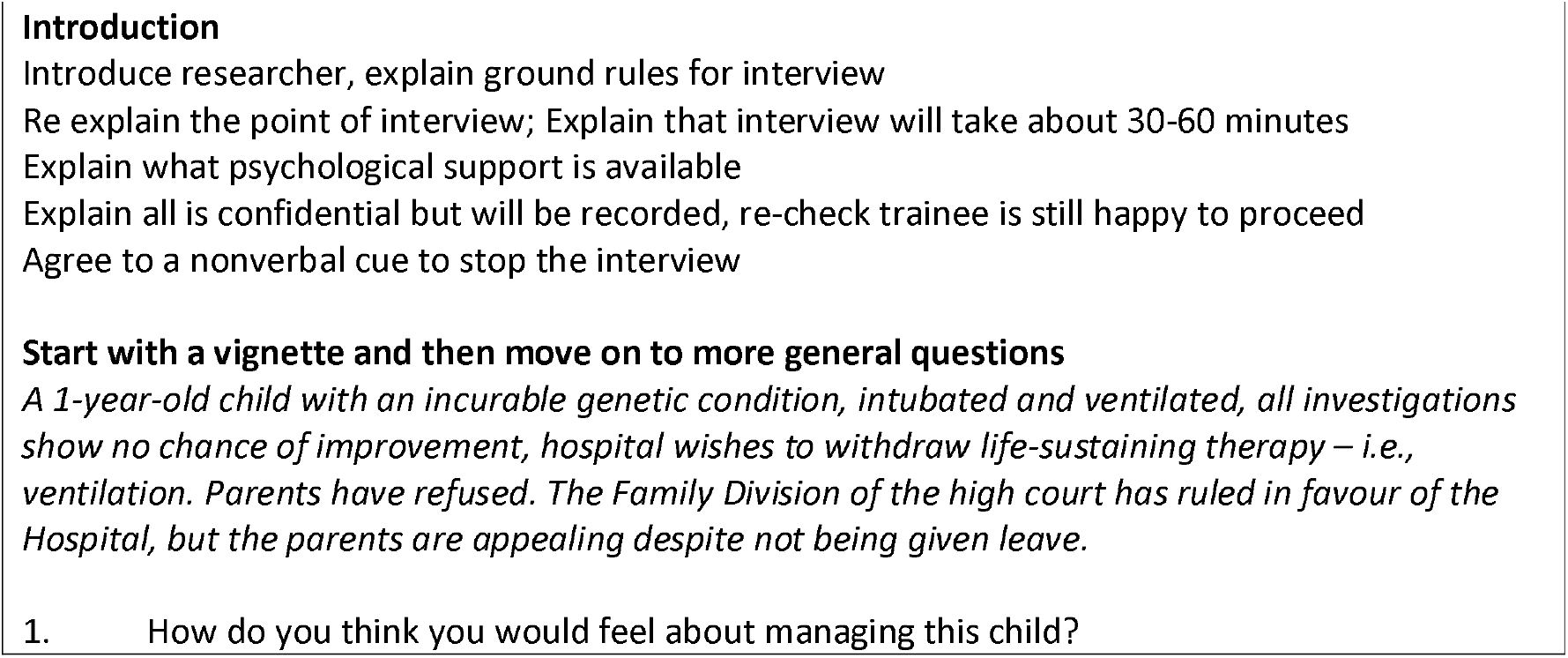
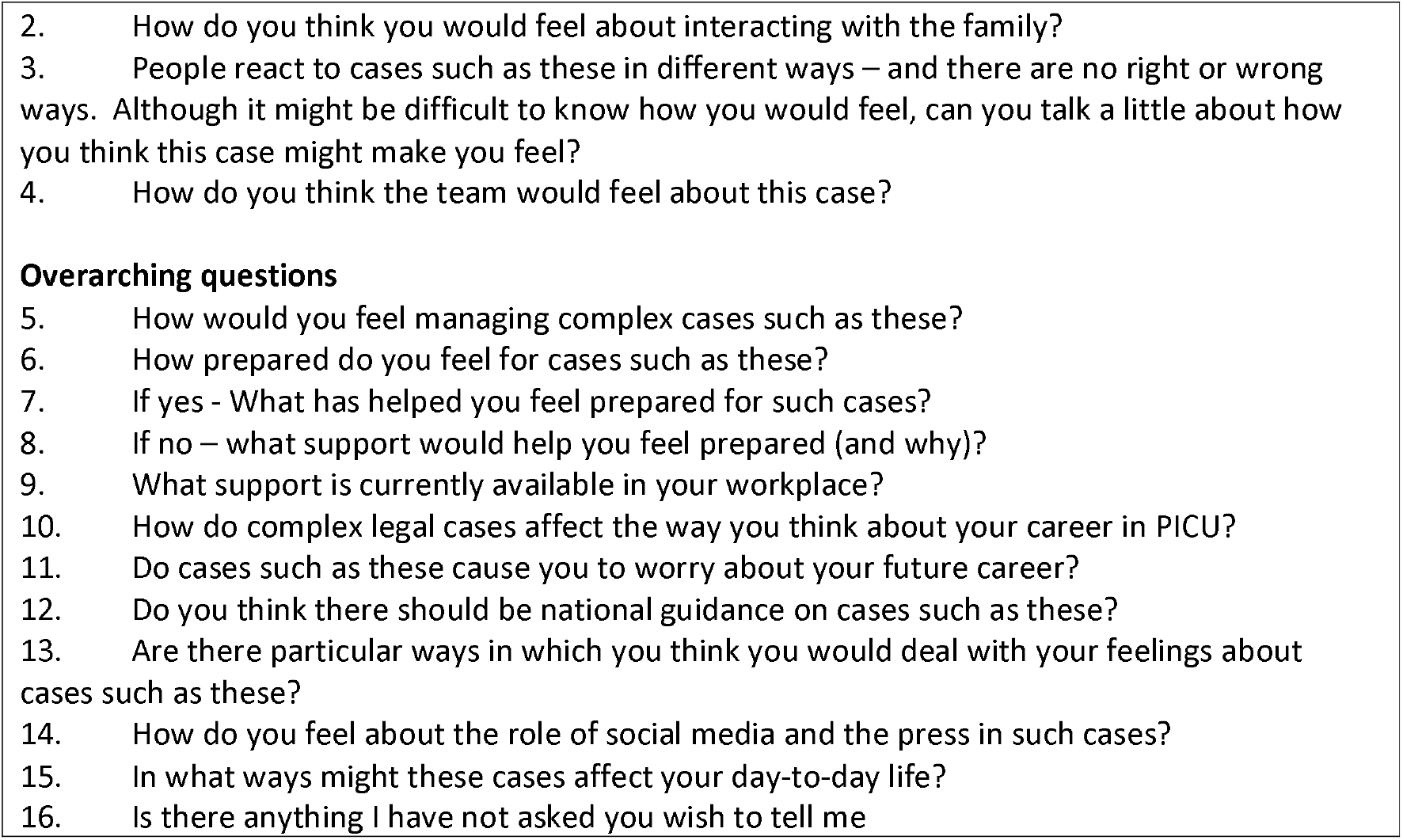
Interview topic guide

**Table 2.** Participant’s demographics (n=9)

| Table 2. Participant's demographics (n=9) |  |
| --- | --- |
|  | Number (n) |
| **Sex** |  |
| Male | 4 |
| Female | 5 |
| **Age range (years)** |  |
| 20-30 | 1 |
| 30-40 | 8 |
| **Geographical area** |  |
| Northern England | 2 |
| South West | 1 |
| London | 3 |
| East of England | 2 |
| South East | 1 |
| **Level of training** |  |
| ST6 | 2 |
| ST7 | 3 |
| ST8 | 4 |
| ST: Specialty Trainee |  |

**Table 2.**

| Theme | Quote |
| --- | --- |
| **Increased specific communication training and teaching in legal and ethics topics are required.** | “The skill that you really can’t do without is you must be able to communicate. If you can’t communicate effectively and you can’t manage difficult situations with families in terms of care consulting, then you really aren’t cut out for the role” (I1) |
|  | “I think communication is the big issue and I like to think I have good communication skills. I’ve done about conflict training and that sort of thing, and I hope that some of those things help me. It will help me in those difficult situations.” (I7) |
|  | “I haven’t seen or heard any controversial or end of life discussions from my seniors, so I have no experience as to how to go about getting to court or what happens when to get to the point where you end up in court” (I8) |
|  | “The fact that they’ve got ethics committees and it probably needs, we need to as trainees be encouraged more or pushing, make ourselves go to more of those ethics committees and that sort of thing” (I2) |
|  | “I am lucky to work with a group of really experienced consultants who do take communication really seriously and has very well demonstrated how to work as a team and how to manage these difficult situations” (I1) |
| **Every case is unique, but more explicit guidance is needed to support PIC doctors, given the number of cases going to court.** | “So having a blanket way of doing it would seem, to me, a little bit insensitive and probably not very professional” (I3) |
|  | “I think as long as you have a good legal team and ethical team to ask for advice and ideally an experienced consultant within your team you wouldn’t need a national guidance. Without a good ethical and legal team or didn’t have an experienced consultant body I feel a national guidance could be useful.” (I8) |
|  | So, I think having some sort of institutionalised set guidelines would be helpful in navigating ethically difficult waters to try and avoid those sorts of conflicts and those sorts of wildly diverging second opinions that might be confusing.” (I1) |
|  | “Would be helpful because once it becomes more protocolised you feel that your personal decisions that you are making as a clinician are at least backed up by a body of evidence or some sort of group decision-making process that isn’t just your own personal bias coming through.” (I1) |

**Table 3.**

| Theme | Quote |
| --- | --- |
| **Care of the child is always the priority, even if it makes trainees feel conflicted.** | ‘I think it’s really important to remember you’re trying to look after the patient and to do the best for the patient regardless of what you, the hospital, and the parents think. As long as I knew what I was doing what was best for the child.’ (I8) |
|  | ‘I would find it hard talking to parents on the ward round and worry about them, I can see with me I’ll be having discussions to going round and round in circles.’ (Interview 8) |
|  | ‘Without doubt the parents are the best advocates for the child in, you know there, there are rare cases where that’s not the case, but I think by and large that is my belief that parents are the best advocates for the child and. Yes, I think I would find that difficult.’ (I2) |
|  | “Your role is there to do what’s the best for the child, it is the doctor’s role to help the child, they are the priority”. (I2) |
|  | “So, it does go contrary to your you know your generic training and your generic practice to then say that this that what, what the parents are saying is not right. So, I think, I think, I think everybody probably struggles with this. I don’t. I don’t think it, it comes naturally to anybody” (I2). |
|  | “I will worry that on the ward this will be the patient people avoid working with, the difficult discussions that parents will start and the parents so I think people would choose not to work with the families.” (I8) |
|  | ‘So, I would, I would do it because we all have to do it, but I would certainly not do more than two days in a row.’ (I3) |
| **Trainees worry about high-profile end-of-life cases and feel unprepared for them.** | “The fact that I saw all these cases made me realise that when I become a consultant that I am going to be expected to deal with these situations. And I felt very inadequate, and I felt very, lost.’ (I2) |
|  | “I think it’d be some, probably one of the main things I would worry about being consultant” (I7) |
|  | “Just thinking about cases like this when you are a consultant, they make you worry about becoming consultant?” (Interview 6) |
|  | ‘So, you don’t get you don’t take part in the ethical committees or committee meetings. You’re you are sort of shielded, which I get. You know, I get why the consultants want to shield you and protect you as trainees. And that is important. It is important to protect the trainees but equally until you expose yourself in these situations. You will never get the motivation to find out about... You know; about all these ethical decisions and about all of that stuff.’ (I2) |
|  | ‘I think I would probably take it home; I take work home with me to a moderate amount already so I imagine I probably wouldn’t just be able to leave it at work. I think it would have reasonably far-reaching consequences.’ (I1) |
|  | “So, yes, I was certainly aware of all the difficult legal cases and what that might look like in practice and how potentially difficult that would be. So, I thought about it, but it did not put me off.” |
|  | “Yeah, I disagree highly when people say it’s okay, this is what you signed up for. This is what is expected. And it might be what’s experienced it’s not what’s expected and that doesn’t mean it’s correct.” (I8) |

**Table 5.**
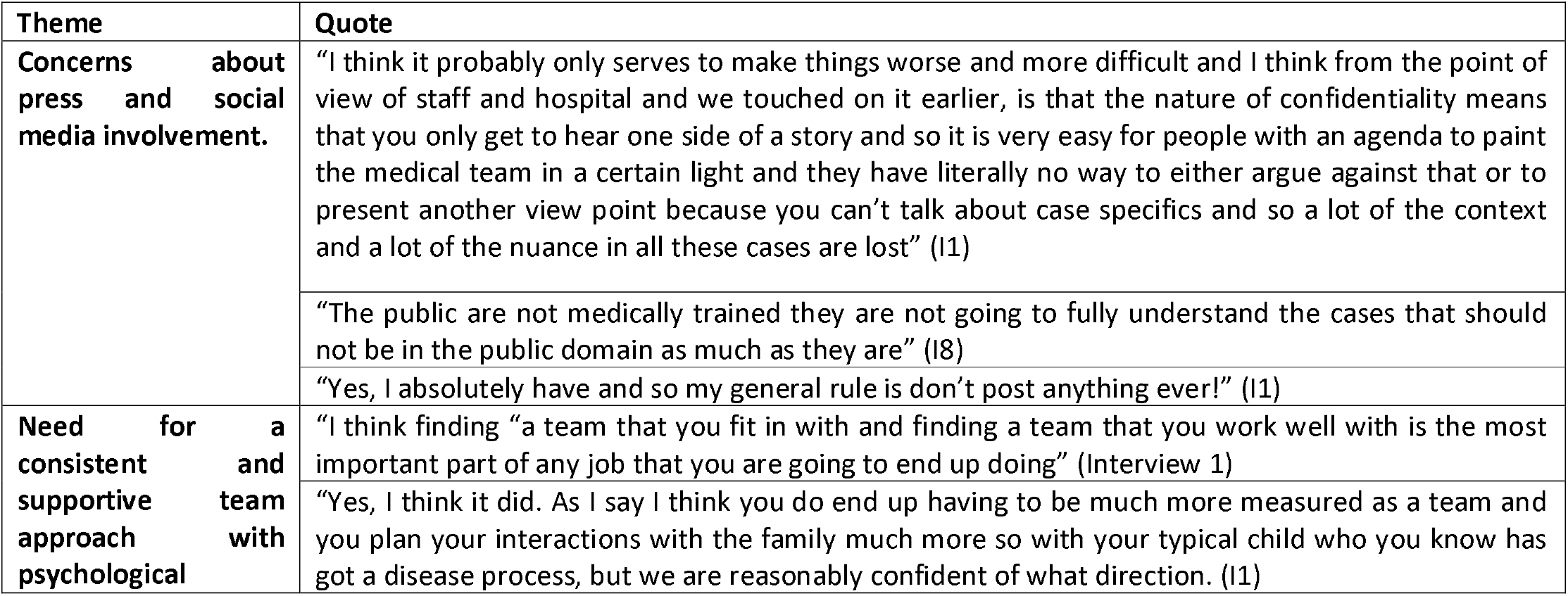
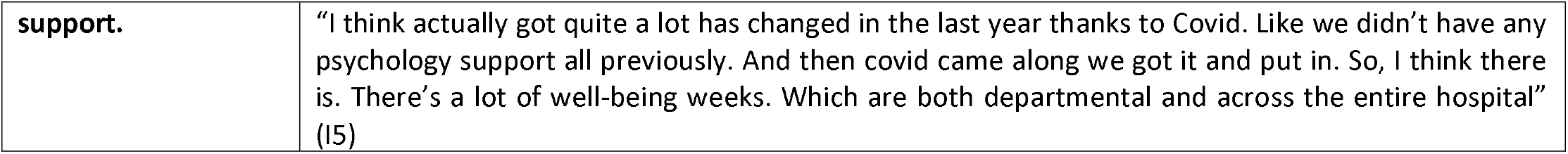

#### 1. Care of the child is always the priority, even if it makes trainees feel conflicted

An overriding theme throughout all interviews, trainees’ priority was doing their best for the child. However, they acknowledge feeling conflicted when disagreeing with parents, making them very uncomfortable. They recognised that parents were the child’s usual best advocate but worried about the harm caused to children when they acceded to non-medically indicated treatment. Despite this, they found it hard to go against parental wishes in practice. In addition, they worried that PIC staff start to avoid looking after some children in these situations, finding caring for them during disagreements strenuous. Concerns were expressed that this causes a lack of continuity of care, with no one wanting to care for the child for more than a couple of days.

#### 2. Trainees worry about high-profile end-of-life cases and feel unprepared for them

All participants expressed serious concerns about managing high-profile end-of-life cases in the future. Participants recognised the responsibility for this rests with PIC consultants but felt vulnerable, unprepared, and too inexperienced to fulfil this role. Participants discussed how the level of responsibility is heightened as a consultant and worried it was hard to understand and experience it as a trainee effectively. Trainees expressed how consultants supportively shielded them from difficult discussions, which, whilst protective, was unhelpful in the long term.

Participants were aware of the emotional burden of these cases on PIC consultants and how the stress involved would be impossible to leave at work, seeping into home life. Furthermore, they were mindful that these cases would be inevitable in the future and take a severe toll on their well-being. Even at this early stage of their career, they foresaw a potential need to step back from clinical duties and seek professional help.

All interviewees thought about such cases seriously when deciding on their careers, considering them a significant downside. Some trainees recognised - but disagreed with - the long-standing belief that in medicine, professionals should just cope with whatever is thrown at you.

#### 3. Increased specific communication training and teaching in legal and ethics topics are required

All interviewees recognised communication skills as an essential skill, especially in challenging cases but perceived it as a potential weakness in their practice. They were clear that observing, practising, and participating in difficult conversations would help prepare them for the consultant role. Ideally, they thought this should be facilitated at work, but alternatively, a dedicated course could be beneficial, though few had attended one.

Additionally, all interviewees wanted to observe both legal and ethics teams directly. They expressed concern that PIC consultants often had discussions with these groups ‘behind closed doors’, preventing them from observing and learning. Many had taken action to fill this gap, 5/9 having sought extracurricular ethics/law education, 2 with MAs in ethics/law.

Many participants highlighted learning from experienced consultants and how they had observed and discussed difficult conversations. In most UK-PICs, complex or long-stay children have a named consultant, and some suggested a named trainee similarly might follow the child’s path and learn the process, though this might be difficult given on-call rotas.

#### 4. Every case is unique, but more explicit guidance is needed to support PIC doctors, given the number of cases going to court

Many trainees felt that high-profile end-of-life cases would continue to increase. However, as each patient is unique, they recognise the difficulties in planning specific treatment or thinking about communication in advance - a bespoke plan and management style are required in each case.

When asked if they thought national guidance and support in controversial end-of-life disputes would be helpful, all agreed with several caveats, depending on the level of preparation and experience of the staff involved. Although several trainees talked about the usefulness of a protocolled set of guidelines that teams could refer to when dealing with parents who disagreed with decisions, a few considered some units better prepared than others, so they thought the usefulness of guidance would vary.

#### 5. Concerns about press and social media involvement

All trainees expressed serious concern about the role of social media and the press in recent high-profile cases; they specifically felt vulnerable due to the lack of anonymity for the teams. Participants expressed frustration at being unable to defend themselves on social media due to patient confidentiality and professional constraints. Finally, participants highlighted how, on social media, people debate issues without being fully informed or understanding the medical facts, so misinformation spreads.

All trainees maintained minimal social media presence due to their jobs and personal feelings about social media. Some did not use their own names on social media, and all were careful about posting content.

#### 6. Need for a consistent and supportive team approach with psychological support

Participants viewed the PIC team as the backbone of their workplace. All highlighted the importance of an empathetic and supportive team with a culture where everyone can talk openly. All trainees felt strongly that the PIC team must be unified in their approach to the child’s care and suggested that each child’s care plan must almost be choreographed. They thought it essential staff have an identical approach and reiterate the same plan, far more for these patients than other groups.

Participants who referred to psychological support noted most was provided informally, but all reported increased well-being support during the Covid-19 pandemic.

## Discussion

The primary aim of this study was to explore the thoughts and feelings of PICU trainees about the impact of recent high-profile end-of-life cases. Their testimonies highlight that all feel unprepared to manage these cases as consultants, and all have significant concerns about the potential emotional distress involved. Whilst this finding was not unexpected, given preceding anecdotal observations, its pervasiveness is concerning. Considering the clear evidence that burnout and moral distress in doctors adversely affects patient care,(11–14) providing PIC trainees with appropriate training and support to deal with high-profile end-of-life cases is vital to ensure children and families receive appropriate care.

Notably, the theme of caring for the child as a priority was paramount. However, there was tension between wanting to do what was best for the child if it conflicted with the parents’ wishes. This tension is mirrored in the literature suggesting most conflicts arise in relation to communication breakdown, mismatching expectations or unrealistic expectations.(15,16)

Trainees universally felt unprepared to manage these cases and worried about the associated emotional burdens, findings comparable to junior staff in child-protection cases.(17) All reflected a need for more teaching and greater involvement in real case discussions, findings consistent with broader European paediatric trainees’ views.(18) The efficacy of introducing this is unknown, so formal assessment would be mandatory. All trainees considered communication skills the cornerstone of managing these cases and wanted opportunities to hone and develop their own, specifically by observing PIC consultants having difficult discussions. They specifically suggested that shadowing hospital ethics and legal teams might equip them with essential skills. A Stanford paediatric palliative care training programme to address deficiencies in doctors’ competencies demonstrated clear improvement in knowledge and confidence. (27) Some participants suggested similar models.

Participants recognised that although high-profile end-of-life cases were increasing in number, each was unique, possibly with common structural failings in management and a changing PIC patient profile (prolonged and recurrent admissions for children with complexity). This paradigm has been seen previously in high-profile child protection cases such as Baby P and Victoria Climbié, with common structural failings but distinct features. (28)

All felt that national guidance about high-profile end-of-life cases would be helpful but must be factual and not replicate existing guidance; some suggested their smaller PIC units might particularly benefit. A national framework for child protection followed the above tragic child deaths, with government inquiries providing structural recommendations. (29) Creating national guidance for high-profile legal cases, or at least undertaking further research on its utility, would seem sensible.

Whilst the GMC and BMA offer guidance for doctors about social media presence (19,20), it is a fast-moving field, and guidelines are out of date almost as soon as release. Despite this, guidance is rarely updated despite clinicians needing accurate, contemporary advice.

There seems little in such guidance, or elsewhere, to advise doctors on what to do if their name is released in the press or on social media in a high-profile end-of-life case. Professional bodies should address this specific issue urgently, given the unacceptable treatment, including death threats, received by clinicians involved in recent cases.(23). Given this, trainees’ feelings of vulnerability and concern about inadequate protection are unsurprising.

In 2005, a Royal College of Physicians Working Party noted that a supportive team, including broader hospital management, was critical for doctors to thrive.(22) Participants in our study recognised that having multi-disciplinary team plans was beneficial to prevent and minimise conflicts and emotional burdens. Pleasingly, all trainees reported that psychological support had improved since the beginning of the COVID-19 pandemic.(23,24)

### Strengths and limitations

Whilst the number interviewed was small, it does represent over 10% of UK PIC trainees. The trainees are a homogenous group in terms of professional background and experience. Due to the Covid-19 pandemic, interviews were conducted virtually, limiting the ability to take notes about non-verbal cues and body language, which could have enriched data interpretation. However, remote interviews allowed for a broad geographical representation within the UK. Data saturation and independent code checking strengthens the validity of our findings. The pandemic was likely to have affected participants’ personal and professional lives, such as unusually stressful work demands and newly established psychological support.

### Future implications and research

Trainee doctors are a sub-section of the multi-disciplinary PIC workforce. It is unlikely all PIC staff have the same views, offering an opportunity to investigate further. Furthermore, formal exploration of the thoughts, feelings and experiences of the families involved in PIC high-profile EOL cases and other stakeholders would be interesting.

## Conclusions

PIC trainees had serious concerns and perceived themselves as inadequate to manage future high-profile end-of-life cases. Our findings demonstrated an unmet training, with participants wanting (i) opportunities to observe and be involved in end-of-life cases, (ii) to strengthen their communication skills and (iii) to both experience and receive formal teaching in ethics and law. PIC training programmes need to address these needs.

More experiential research in this area, specifically with the families involved and other key stakeholders, would complete a rounded picture of the current situation.

### Contributory statement

CB and JB conceived the presented idea, CB and JB designed the study and CB completed the interviews. CB, JB and MD analysed the data. CB, JB and MD wrote the manuscript.

## Supporting information

COREQ Q

PIP

## Data Availability

Deidentified participant interview transcripts, are available from Dr Clare Bell, ORCID identifier 0000-0002-8331-5758 reuse is permitted after discussion with the research team in related research.

## Acknowledgements

Thank you to Professor Helen Bedford and Professor Paul Winyard.

## References

1. Tume LN, Menzies JC, Ray S, Scholefield BR. Research Priorities for U.K. Pediatric Critical Care in 2019: Healthcare Professionals’ and Parents’ Perspectives. Pediatr Crit care Med a J Soc Crit Care Med World Fed Pediatr Intensive Crit Care Soc. 2021 May;22(5):e294–301.

2. Prentice T, Janvier A, Gillam L, Davis PG. Moral distress within neonatal and paediatric intensive care units: a systematic review. Arch Dis Child. 2016;101(8):701–8.

3. Rodríguez-Rey R, Palacios A, Alonso-Tapia J, Pérez E, Álvarez E, Coca A, et al. Burnout and posttraumatic stress in paediatric critical care personnel: Prediction from resilience and coping styles. Aust Crit care Off J Confed Aust Crit Care Nurses. 2019 Jan;32(1):46–53.

4. Hurley R. How a fight for Charlie Gard became a fight against the state. BMJ. 2017 Aug;358:j3675.

5. Dyer C. Children’s hospital must be allowed to withdraw life support from Alfie Evans, court rules. BMJ. 2018;361.

6. Dyer C. Hospital trust asks High Court to rule whether it can withdraw girl’s life support. BMJ. 2019;366.

7. Cass H, Barclay S, Gerada C, Lumsden DE, Sritharan K. Complexity and challenge in paediatrics: a roadmap for supporting clinical staff and families. Arch Dis Child. 2020;105(2):109–14.

8. Jones GAL, Colville GA, Ramnarayan P, Woolfall K, Heward Y, Morrison R, et al. Psychological impact of working in paediatric intensive care. A UK-wide prevalence study. Arch Dis Child. 2020 May;105(5):470–5.

9. Schwandt TA. Dictionary of qualitative inquiry. Second edition. Thousand Oaks, Calif.l1: Sage Publications, [2001] ©2001;

10. Braun V, Clarke V. Using thematic analysis in psychology. Qual Res Psychol. 2006;3(2):77–101.

11. Kumar S. Burnout and Doctors: Prevalence, Prevention and Intervention. Healthc (Basel, Switzerland). 2016 Jun;4(3).

12. Lerkiatbundit S, Borry P. Moral Distress Part I: Critical Literature Review on Definition, Magnitude, Antecedents and Consequences. J Pharm Pract. 2009;1(1).

13. Steel K. Geriatrics-A Profession in the Making. Journals Gerontol Ser A Biol Sci Med Sci. 2004 Nov;59(11):1168–9.

14. Hodkinson A, Zhou A, Johnson J, Geraghty K, Riley R, Zhou A, et al. Associations of physician burnout with career engagement and quality of patient care: systematic review and meta-analysis. BMJ. 2022;378.

15. Forbat L, Sayer C, McNamee P, Menson E, Barclay S. Conflict in a paediatric hospital: a prospective mixed-method study. Arch Dis Child. 2016;101(1):23–7.

16. François K, Lobb E, Barclay S, Forbat L. The nature of conflict in palliative care: A qualitative exploration of the experiences of staff and family members. Patient Educ Couns. 2017;100(8):1459–65.

17. Ashtekar CS, Hande A, Stallard E, Tuthill D. How much do junior staff know about common legal situations in paediatrics? Child Care Health Dev. 2007 Sep;33(5):631–4.

18. Boer MC den, Zanin A, Latour JM, Brierley J. Paediatric Residents and Fellows Ethics (PERFEct) survey: perceptions of European trainees regarding ethical dilemmas. Eur J Pediatr. 2022;181(2):561–70.

19. British Medical Association. Social media, ethics and professionalism: BMA guidance. London Br Med Assoc. 2018;

20. General Medical Council. Doctors’ use of social media. Gen Med Counc. 2013;(April):1–4.

21. Lagercrantz H. Observations on the case of Charlie Gard. Arch Dis Child. 2018;103(5):409–10.

22. Doctors in society. Medical professionalism in a changing world. Clin Med. 2005;5(6 Suppl 1):S5–40.

23. Behrman S, Baruch N, Stegen G. Peer support for junior doctors: a positive outcome of the COVID-19 pandemic? Futur Healthc J. 2020 Oct;7(3):e64–6.

24. Faderani R, Monks M, Peprah D, Colori A, Allen L, Amphlett A, et al. Improving wellbeing among UK doctors redeployed during the COVID-19 pandemic. Futur Healthc J. 2020;7(3):e71–6.

