## Supplementary material for "What is the impact of high-profile end-of-life disputes on Paediatric Intensive Care (PIC) trainees? Original research": COREQ Q

**Table 1**

Consolidated criteria for reporting qualitative studies (COREQ): 32-item checklist

| **No** | **Item** | **Guide questions/description** | **Action** | **Page** |
| --- | --- | --- | --- | --- |
| **Domain 1: Research team and reflexivity** |  |  |  |  |
| Personal Characteristics |  |  |  |  |
| 1. | Interviewer/facilitator | Which author/s conducted the interview or focus group? | CB – Paediatric UK ST8 final year trainee | 5 |
| 2. | Credentials | What were the researcher's credentials? *E.g. PhD, MD* | CB – MBBS, BSc, MSc | 5 |
| 3. | Occupation | What was their occupation at the time of the study? | Paediatric Registrar | 5 |
| 4. | Gender | Was the researcher male or female? | Female | 5 |
| 5. | Experience and training | What experience or training did the researcher have? | MSc Advanced paediatrics UCL, UCL/GOSH Qualitative research 5 day course | 5 |
| Relationship with participants |  |  |  |  |
| 6. | Relationship established | Was a relationship established prior to study commencement? | CB had worked 7 years previously with 1 trainee for 6 months | 5 |
| 7. | Participant knowledge of the interviewer | What did the participants know about the researcher? e*.g. personal goals, reasons for doing the research* | Trainees were told of CBs job and sent a PIP explaining the background to the research | In supplemental info |
|  | Interviewer characteristics | What characteristics were reported about the interviewer/facilitator? e.g. *Bias, assumptions, reasons and interests in the research topic* | Bias and reflexivity noted in methods | 5 |
| **Domain 2: study design** |  |  |  |  |
| Theoretical framework |  |  |  |  |
| 9. | Methodological orientation and Theory | What methodological orientation was stated to underpin the study? *e.g. grounded theory, discourse analysis, ethnography, phenomenology, content analysis* | Thematic analysis |  |
| Participant selection |  |  |  |  |
| 10. | Sampling | How were participants selected? *e.g. purposive, convenience, consecutive, snowball* | Purposeful sampling | 5 |
| 11. | Method of approach | How were participants approached? e*.g. face-to-face, telephone, mail, email* | Email via a third party | 5 |
| 12. | Sample size | How many participants were in the study? | 9 participants | 5 |
| 13. | Non-participation | How many people refused to participate or dropped out? Reasons? | No one dropped out, 1 trainee did not meet the inclusion criteria | 5 |
| Setting |  |  |  |  |
| 14. | Setting of data collection | Where was the data collected? e*.g. home, clinic, workplace* | Data collected at workplace (GOSH) via remote interviews | 5 |
| 15. | Presence of non-participants | Was anyone else present besides the participants and researchers? | no | 5 |
| 16. | Description of sample | What are the important characteristics of the sample? *e.g. demographic data, date* | In table 2 | 7 and 15 |
| Data collection |  |  |  |  |
| 17. | Interview guide | Were questions, prompts, guides provided by the authors? Was it pilot tested? | Was pilot tested, prompts were used, as seen in addendum | 5 |
| 18. | Repeat interviews | Were repeat interviews carried out? If yes, how many? | no |  |
| 19. | Audio/visual recording | Did the research use audio or visual recording to collect the data? | Audio recording | 6 |
| 20. | Field notes | Were field notes made during and/or after the interview or focus group? | yes | 6 |
| 21. | Duration | What was the duration of the interviews or focus group? | 30-90 min | 6 |
| 22. | Data saturation | Was data saturation discussed? | yes | 13 |
| 23. | Transcripts returned | Were transcripts returned to participants for comment and/or correction? | no | 6 |
| **Domain 3: analysis and findings** |  |  |  |  |
| Data analysis |  |  |  |  |
| 24. | Number of data coders | How many data coders coded the data? | 1 | 6 |
| 25. | Description of the coding tree | Did authors provide a description of the coding tree? | yes | 6 |
| 26. | Derivation of themes | Were themes identified in advance or derived from the data? | Derived from the data | 6 |
| 27. | Software | What software, if applicable, was used to manage the data? | NVivo® | 6 |
| 28. | Participant checking | Did participants provide feedback on the findings? | no | 6 |
| Reporting |  |  |  |  |
| 29. | Quotations presented | Were participant quotations presented to illustrate the themes / findings? Was each quotation identified? e*.g. participant number* | yes | 7-11 |
| 30. | Data and findings consistent | Was there consistency between the data presented and the findings? | yes | 7-11 |
| 31. | Clarity of major themes | Were major themes clearly presented in the findings? | yes | 7-11 |
| 32. | Clarity of minor themes | Is there a description of diverse cases or discussion of minor themes | yes | 7-11 |
