## Supplementary material for "What is the impact of high-profile end-of-life disputes on Paediatric Intensive Care (PIC) trainees? Original research": PIP

**Addendum**

**Participant Information Sheet.**

**Study Title** - End of life high profile court battles - Study of views and feelings of Paediatric Intensive care trainees?

Version 1.0, IRAS ID

**1. An invitation**

We would like to invite you to take part in a research project. This will involve taking part in a 30-60 minute interview about your feelings and views on high profile end of life court cases.

**2. What is the purpose of the study?**

The purpose of the study is to understand if and how the high profile cases have affected PICU trainees, and if they have to what extent. When this is understood is there is a detrimental effect on trainees mechanisms can they be put in place to lessen the negative effects

**3. Why have I been invited to take part?**

You have been invited to take part because you are PICU GRID trainee.

**4. Do I have to take part?**

It is up to you to decide whether to take part or not. You will be given time to go through this information leaflet and the opportunity to ask questions. If you decide to take part, you are free to withdraw at any time without giving a reason.

**5. What will happen to me if I decide to take part?**

You will email the researcher – Dr Clare Bell and she will arrange an interview either in person at time of your choosing. The interview will last approximately 30-60 minutes. All discussed is totally confidential but direct non attributable quotes maybe used in the completion of the research. The interview will be recorded on an encrypted secure devise.

**6. What are the possible disadvantages or risks of taking part?**

For this research study, the main disadvantages are that you will be asked to give up some of your time and discuss potentially emotive topics.

**7. What are the possible benefits of taking part?**

You will be helping understanding how PICU trainees feel about these cases

**8. Will my part in this study be kept confidential?**

University College London- Institute of Child Health is the sponsor for this study based in the United Kingdom. We will be using information from you in order to undertake this study and will act as the data controller for this study. This means that we are responsible for looking after your information and using it properly. UCL-ICH will keep identifiable information about you for 1 year.

Your rights to access, change or move your information are limited, as we need to manage your information in specific ways in order for the research to be reliable and accurate. If you withdraw from the study, we will keep the information about you that we have already obtained. To safeguard your rights, we will use the minimum personally-identifiable information possible.

You can find out about how we use your information by contacting Dr Clare Bell.

**9. What happens after the study stops?**

After the study stops, you will be emailed a summary of the research findings and told how you can access the information resource, if requested. You will not be contacted again by the researcher.

**10. What if there is a problem?**

If you have a concern about any aspect of this study, you should ask to speak with the researcher who will do their best to answer your question(s). If you remain unhappy and wish to complain formally, you can do this either through the NHS Complaints Procedure or through the UCL University complaints office. The researchers, or any health care professional, will be able to give you more information about these procedures.

**11. What if I need more support**

If you feel you need more support Dr Clare Bell can provide support, there is also the project supervisor Dr Brierley available and a counsellor if you need to discuss issues further. There is also support in your local hospital from the wellbeing team and your educational supervisor.

**12. What will happen if I don’t want to carry on with the study?**

You are free to withdraw at any time and without giving a reason.

**13. What will happen to the results of this research project?**

The main results of this research will be sent to participants, if requested, after it finishes and will usually be published in a peer reviewed medical journal or be presented at a scientific conference. All the data will be anonymous.

**14. Who is organising study?**

The study is being organised by Dr Clare Bell, the study is sponsored by University College London/Great Ormond Street.

**16. Further information**

You are encouraged to ask any questions you wish, before, during or after your participation. If you have any questions about the study, please speak to Clare.

Version 1, 12/02/2020
